# Sublingual microcirculatory alterations during the immediate and early postoperative period: A systematic review and meta-analysis

**DOI:** 10.1101/2021.04.03.21254867

**Authors:** Athanasios Chalkias, Nikolaos Papagiannakis, Georgios Mavrovounis, Konstantina Kolonia, Maria Mermiri, Ioannis Pantazopoulos, Eleni Laou, Eleni Arnaoutoglou

## Abstract

**Study objective:** To systematically review the literature regarding the presence of sublingual microcirculatory alterations during the immediate and early postoperative period.

**Design:** Systematic review and meta-analysis searching PubMed, Scopus, and Google Scholar.

**Patients/interventions:** Studies comparing sublingual microcirculation before and after surgery.

**Measurement:** The primary outcome was to investigate the severity of microcirculatory alterations during the immediate and early postoperative period in adult patients undergoing non-cardiac and cardiac surgery.

**Main results:** Among 17 eligible studies, 13 were finally analyzed. A non-statistically significant difference was found between preoperative and postoperative total vessel density (p=0.084; estimate: −0.029; 95% CI: −0.31 to 0.26; I^2^ = 22.55%, Q = 10.23, p=0.24). Perfused vessel density significantly decreased postoperatively (p=0.035; estimate: 0.344; 95% CI: 0.02 to 0.66; I^2^ = 65.66%, Q = 41.77, p<0.001), while perfused boundary region significantly increased postoperatively (p=0.031; estimate: −0.415; 95% CI: −0.79 to −0.03; I^2^ = 37.21%, Q = 6.56, p=0.16). Microvascular flow index significantly decreased postoperatively (p=0.028; estimate: 0. 587; 95% CI: 0.06 to 1.11; I^2^ = 86.09%, Q = 96.28, p<0.001), while a non-statistically significant difference was found between preoperative and postoperative proportion of perfused vessels (p=0.089; estimate: 0.53; 95% CI: −0.08 to 1.14; I^2^ = 70.71%, Q = 18.99, p=0.002). The results in the non-cardiac surgery subgroup were comparable with the full group except that a statistically non-significant difference in PVD was found in the remaining seven studies (p=0.19; estimate: 0.26; 95% CI: −0.13 to 0.66; I^2^ = 71.94%, Q = 33.42, p=0.002). The other parameters did not differ significantly from their respective full group results.

**Conclusions:** Significant sublingual microcirculatory alterations are present during the immediate and early postoperative period.

## 1. INTRODUCTION

Although the microcirculation has been described since the 17^th^ century, it has gained importance during the last three decades following the introduction of hand-held vital microscopes. Since then, much progress has been made in our understanding of microcirculatory flow and its importance. This terminal vascular network of the systemic circulation is responsible for tissue oxygenation, transport of hormones and nutrients to the tissue cells, and has a central role in mediating the functional activity of the immune system and hemostasis [1].

Microcirculation is highly adaptive to its cellular environment and can be autoregulated, ensuring a constant blood flow that is independent of changes in systemic blood pressure [2]. This hemodynamic coherence is fundamental for the preservation of homeostasis and organ function. However, several conditions, such as sepsis or shock, changes in blood viscosity and shear stress, and iatrogenic injury, may result in an uncoupling between the macro- and microcirculation and tissue injury [3]. This is very important for the perioperative setting considering the diversity of patients, the severity of illness, type of surgery (cardiac *vs*. non-cardiac), and the dynamic intraoperative environment [1]. During the intraoperative period, several parameters (e.g., surgical manipulations, hemorrhage, hypothermia, anesthetics, vasopressors, etc.) may result in microcirculatory alterations and tissue hypoperfusion. Nevertheless, the improvement in systemic hemodynamic parameters does not necessarily result in a parallel improvement in microcirculatory flow.

Despite recent advances, patients undergoing major surgery still suffer from postoperative adverse events and a thorough perioperative evaluation is recommended by several societies to decrease morbidity and mortality. However, the assessment of sublingual microcirculation in this population is not routinely recommended. On the other hand, the incidence and the effects of microcirculatory impairment on postoperative outcome have not been studied extensively. In addition, considering that the derangement of microcirculation occurs in a highly heterogeneous, organ-specific pattern, it is possible that it continues to be underappreciated clinically. Therefore, the objective of this systematic review and meta-analysis was to investigate the presence of sublingual microcirculatory alterations during the immediate and early postoperative period.

## 2. MATERIALS AND METHODS

### 2.1 Protocol and registration

The protocol was registered in the PROSPERO international prospective register of systemic reviews on June 8, 2020 (CRD42020190955). This systematic review and meta-analysis were reported according to the preferred reporting items for systematic reviews and meta-analyses (PRISMA) protocols [4].

### 2.2 Inclusion and exclusion criteria

All randomized controlled trials, clinical trials, comparative studies, cohort studies, pragmatic clinical trials, validation studies, and observational studies were included. Case reports, case series, review papers, and animal studies were excluded. Articles on tissue flaps and other plastic surgical/cosmetic procedures, organ transplantation, and non-English literature were also excluded.

### 2.3 Outcomes of interest

The primary outcome was to investigate the severity of sublingual microcirculatory alterations during the immediate and early postoperative period in adult patients undergoing surgery. Secondary outcome was to investigate the association between postoperative sublingual microcirculatory alterations and complications, admission to the intensive care unit (ICU), and survival following surgery.

### 2.4 Search strategy

The search strategy was intended to explore all available published and unpublished studies from January 2000 to March 2021. A comprehensive initial search was employed in PubMed and Scopus followed by an analysis of the text words contained in Title/Abstract and indexed terms (Supplementary File 1). A second search was conducted by combining free text words and indexed terms with Boolean operators. A third search was conducted with the reference lists of all identified reports and articles for additional studies. Finally, a grey literature search was conducted on Google Scholar.

In the abstract or title, the search contained the MeSH® terms microcirculation, the wildcard terms microcirc*or capilar*, or any of the following terms; microvascular, sublingual, sidestream-dark field imaging, SDF-imaging, microscan, incident dark field imaging, IDF-imaging, cytoscan, orthogonal polarisation imaging, or OPS imaging. Then, the search included articles which meet the above criteria and any of the following MeSH® terms: postoperative, surgery, or surgical or the following non-MeSH terms: complications, outcome, or survival. In addition, the Clinicaltrials.gov website was searched for all articles containing any of the following terms: microcirculation, capillary, microvascular, orthogonal polarisation spectral imaging, sidestream dark-field imaging, and incident dark field imaging.

### 2.5 Data extraction

The data from each study were extracted by two independent authors with a customized format. Any disagreements between the two independent authors were resolved by two other authors. Publication details (authors, year), study information (design, population, follow-up, inclusion-exclusion criteria, and number of cases/cohort-size, subgroups), method/device for assessment of microcirculation, microcirculatory parameters (Supplementary Table 1), hospital length of stay, and outcome were extracted in a pre-designed spreadsheet. Authors of studies with missing data were contacted in an attempt to obtain relevant data.

### 2.6 Assessment of methodological quality

Articles identified for retrieval were assessed by three independent authors for methodological quality before inclusion in the review using a standardized critical appraisal tool. Any disagreements between the authors appraising the articles were resolved through discussion with the other authors.

### 2.7 Data analysis

We applied random-effects meta-analysis to standardized mean differences (SMD) estimating the associations between pre- and postoperative measurements. Random-effects models were a priori preferred over fixed-effects models due to the expected heterogeneity between studies with regards to exposure and outcome definition. The significance level for the overall effect was set at p < 0.05. Between-study heterogeneity was assessed by the I^2^ and the Cochran Q test; significance level was defined by a P value < 0.05 or I^2^ > 50%. Estimation of means and standard deviation in studies reporting medians and interquartile ranges was performed using the techniques described in McGrath et al. [5] Many of the studies reported their microcirculation results by categories and not overall. In order to avoid using two levels of aggregating results, one by study and one by measured quantity, we reported and synthesized separately each result from the different categories of each study.

Due to the limited number of studies included in each analysis, no statistical assessment of publication bias or meta-regression for the association with different study outcomes could be performed. The Cochrane Handbook for systematic reviews, also recommends against it when there are less than 10 studies available [6]. For the same reason, a subgroup analysis was inappropriate. As there was no access to individual data, it was not possible to estimate the correlation between complications and alterations in microcirculation, given the difficulty in performing meta-regression. An exception was made for studies involving cardiac surgery, because these patients frequently require extracorporeal circulation that is clearly associated with microcirculatory alterations. An extra synthesis was performed where studies on cardiac surgery patients were excluded. All analyses were performed using R v. 4.0 (R foundation) with ‘metafor’ and ‘estmeansd’ package.

## 3. RESULTS

Altogether, 14,712 relevant citations were identified and screened, while 17 studies were included in our final assessment for possible data extraction (Fig.1). In total, data extraction was possible in four studies for total vessel density (TVD) [7-10], seven studies for perfused vessel density (PVD) [7-9,11-14], two studies for perfused boundary region (PBR) [11,15], eight studies for microvascular flow index (MFI) [8,9,12-14,16-18], and three for proportion of perfused vessels (PPV) (Supplementary Tables 2-6) [8,12,14].

**Figure 1.**
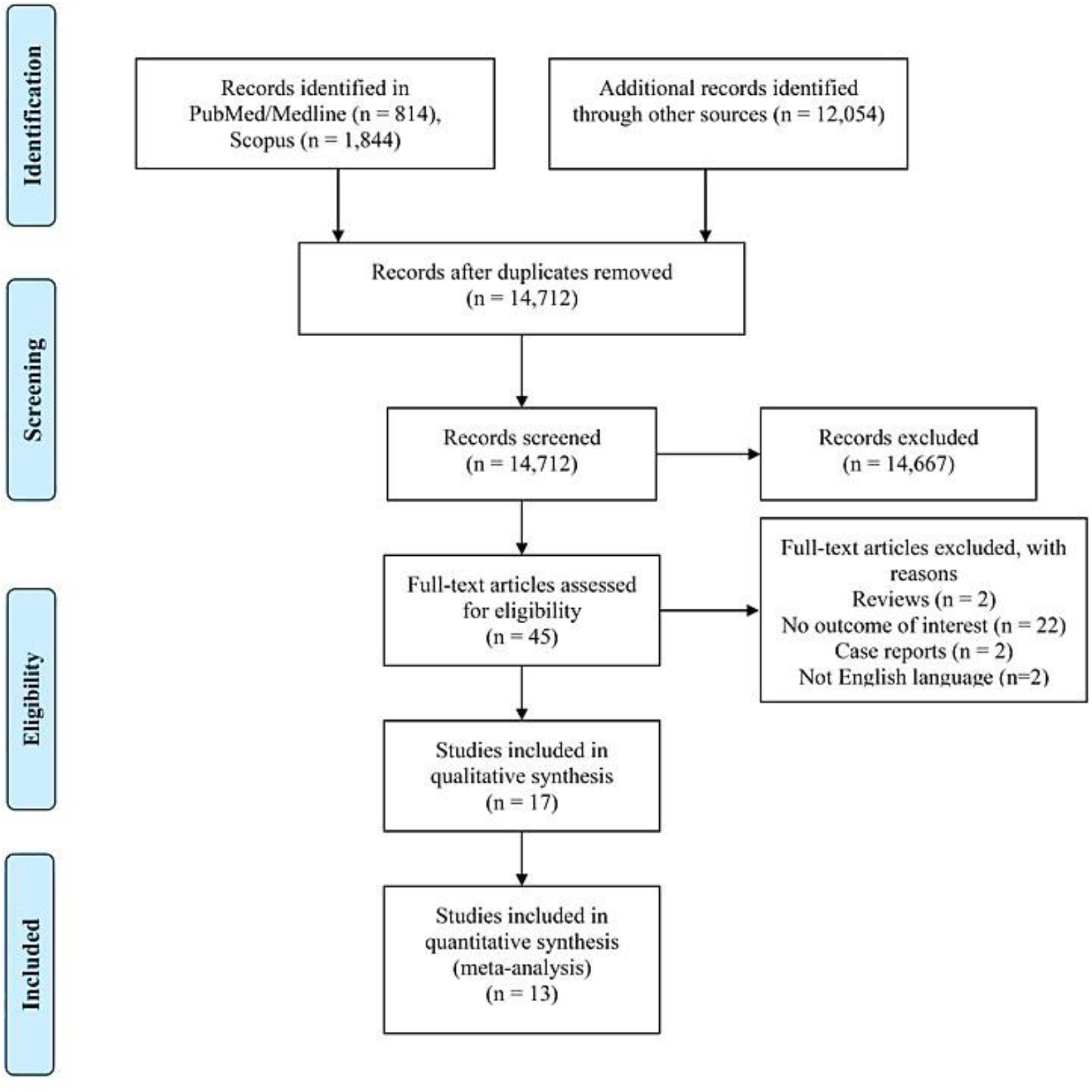
Preferred Reporting Items for Systematic Reviews and Meta-Analyses (PRISMA) diagram.

### 3.1 Synthesis including all patients

A statistically non-significant difference was found between preoperative and postoperative TVD (p=0.084; estimate: −0.029; 95% CI: −0.31 to 0.26). The results were mostly consistent in the four studies so low heterogeneity was present (I^2^ = 22.55%, Q

= 10.23, p=0.24). Perfused vessel density significantly decreased postoperatively (p=0.035; estimate: 0.344; 95% CI: 0.02 to 0.66) with medium to high heterogeneity (I^2^ = 65.66%, Q = 41.77, p<0.001). Perfused boundary region significantly increased postoperatively (p=0.031; estimate: −0.415; 95% CI: −0.79 to −0.03) without the presence of high degrees of heterogeneity (I^2^ = 37.21%, Q = 6.56, p=0.16). Microvascular flow index significantly decreased postoperatively (p=0.028; estimate: 0.587; 95% CI: 0.06 to 1.11) with high degrees of heterogeneity (I^2^ = 86.09%, Q = 96.28, p<0.001). A statistically non-significant difference was found between preoperative and postoperative PPV (p=0.089; estimate: 0.53; 95% CI: −0.08 to 1.14) with high heterogeneity (I^2^ = 70.71%, Q = 18.99, p=0.002). Forest plots for the different measurements are presented in Figures 2-6 and a summary of the results is presented in Supplementary Table 7.

**Figure 2.**
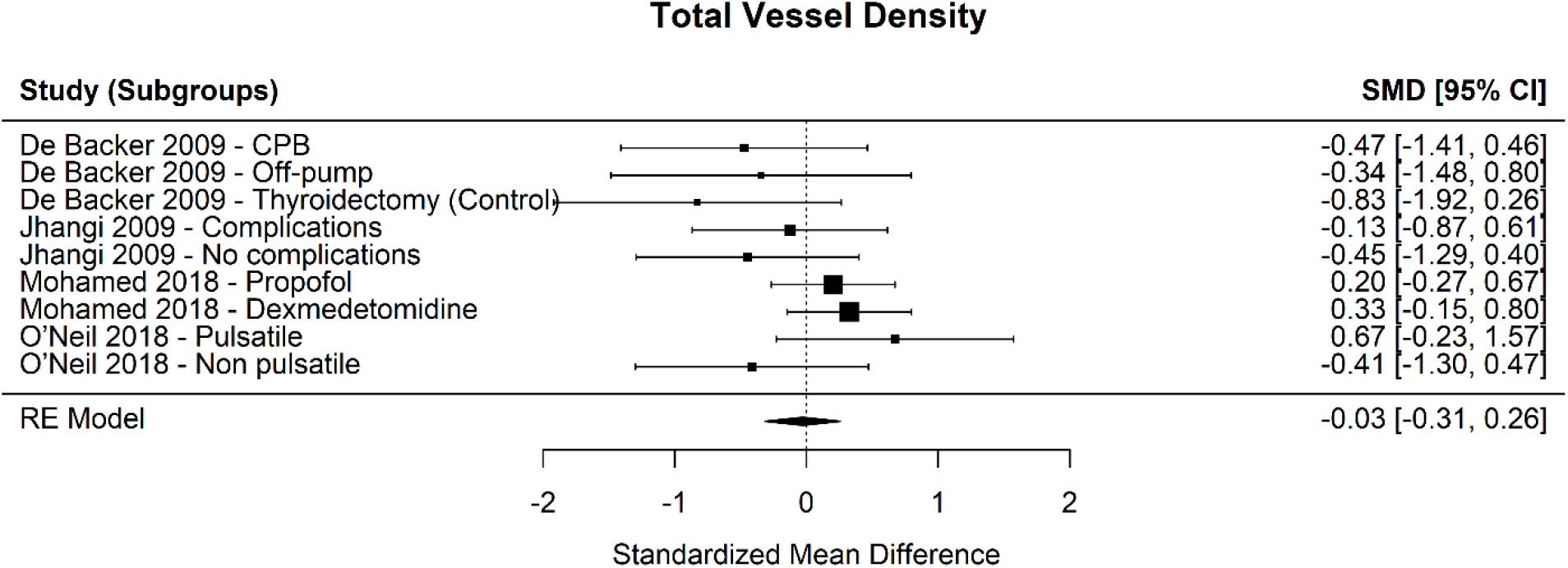
Forest plot for total vessel density. SMD, standardized mean difference; CI, confidence interval.

**Figure 3.**
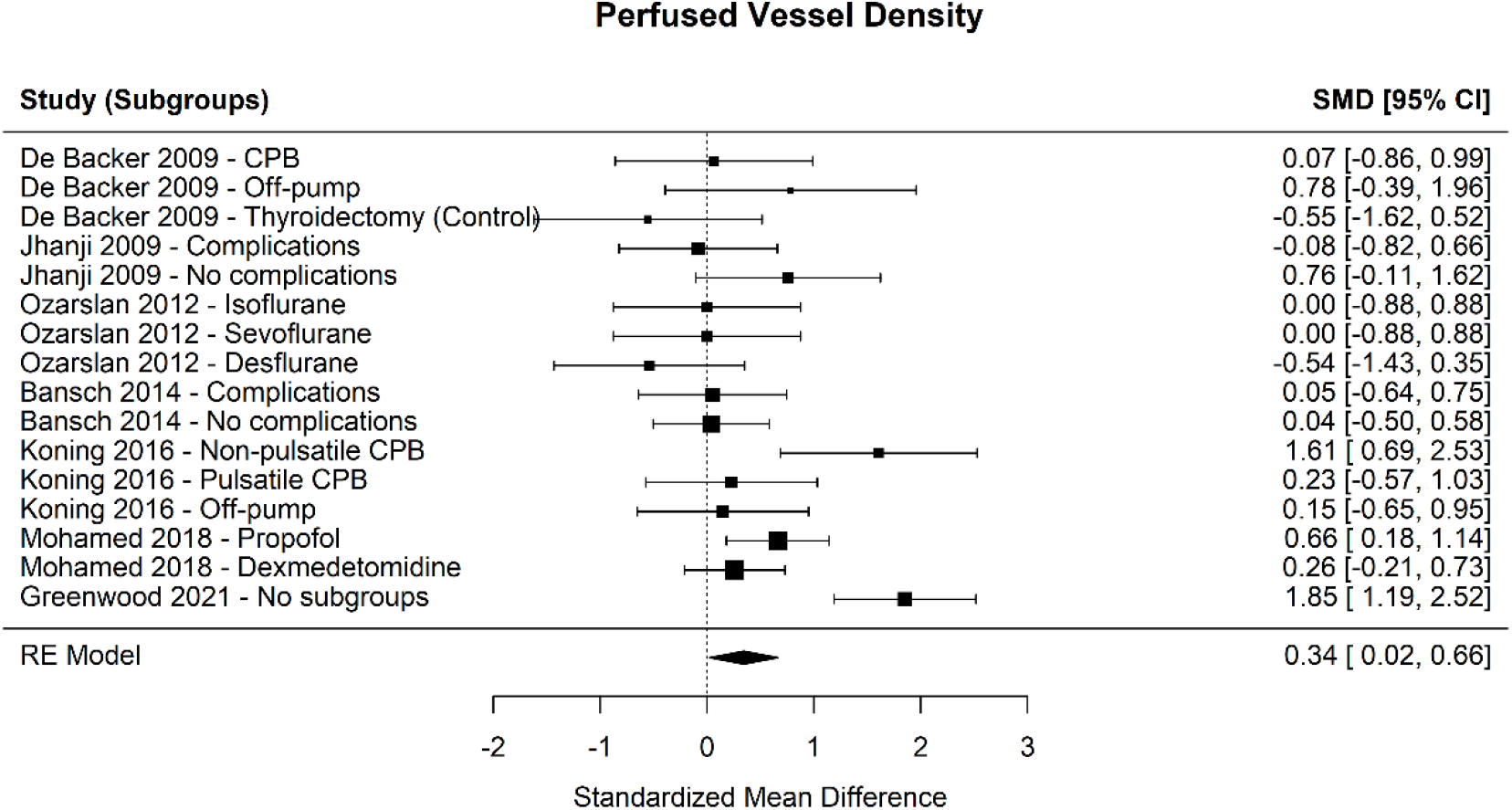
Forest plot for perfused vessel density. SMD, standardized mean difference; CI, confidence interval.

**Figure 4.**
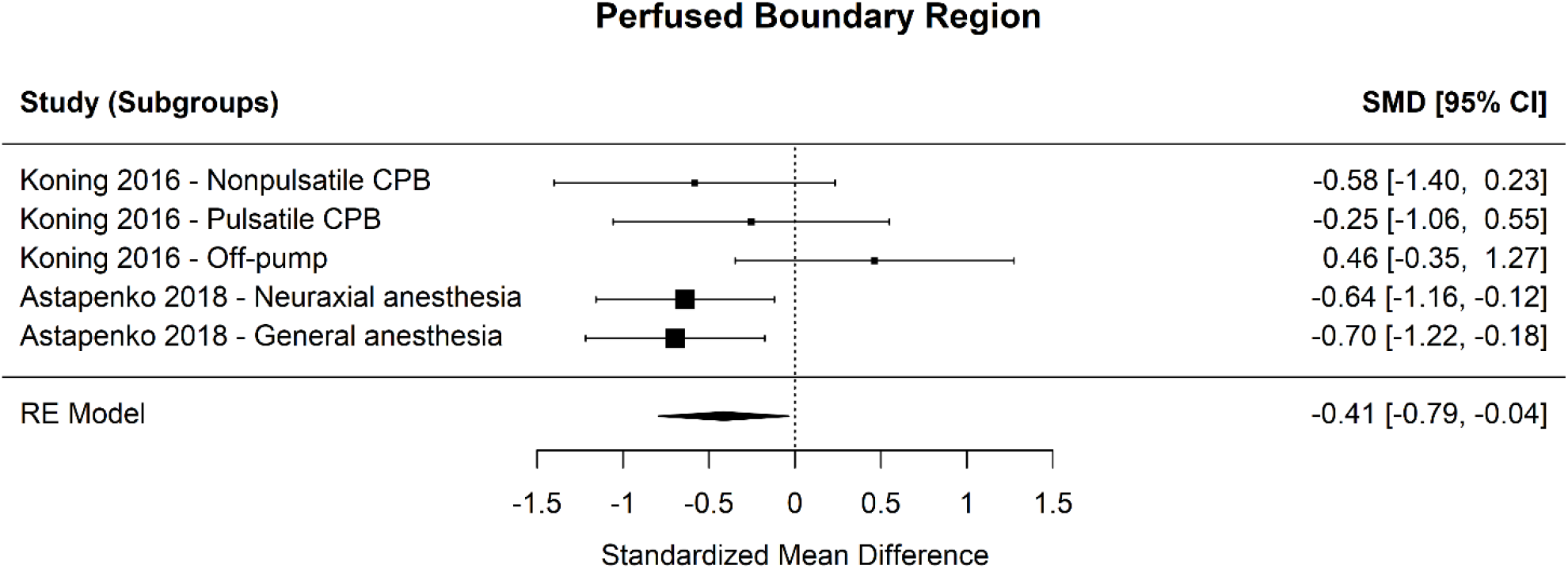
Forest plot for perfused boundary region. SMD, standardized mean difference; CI, confidence interval.

**Figure 5.**
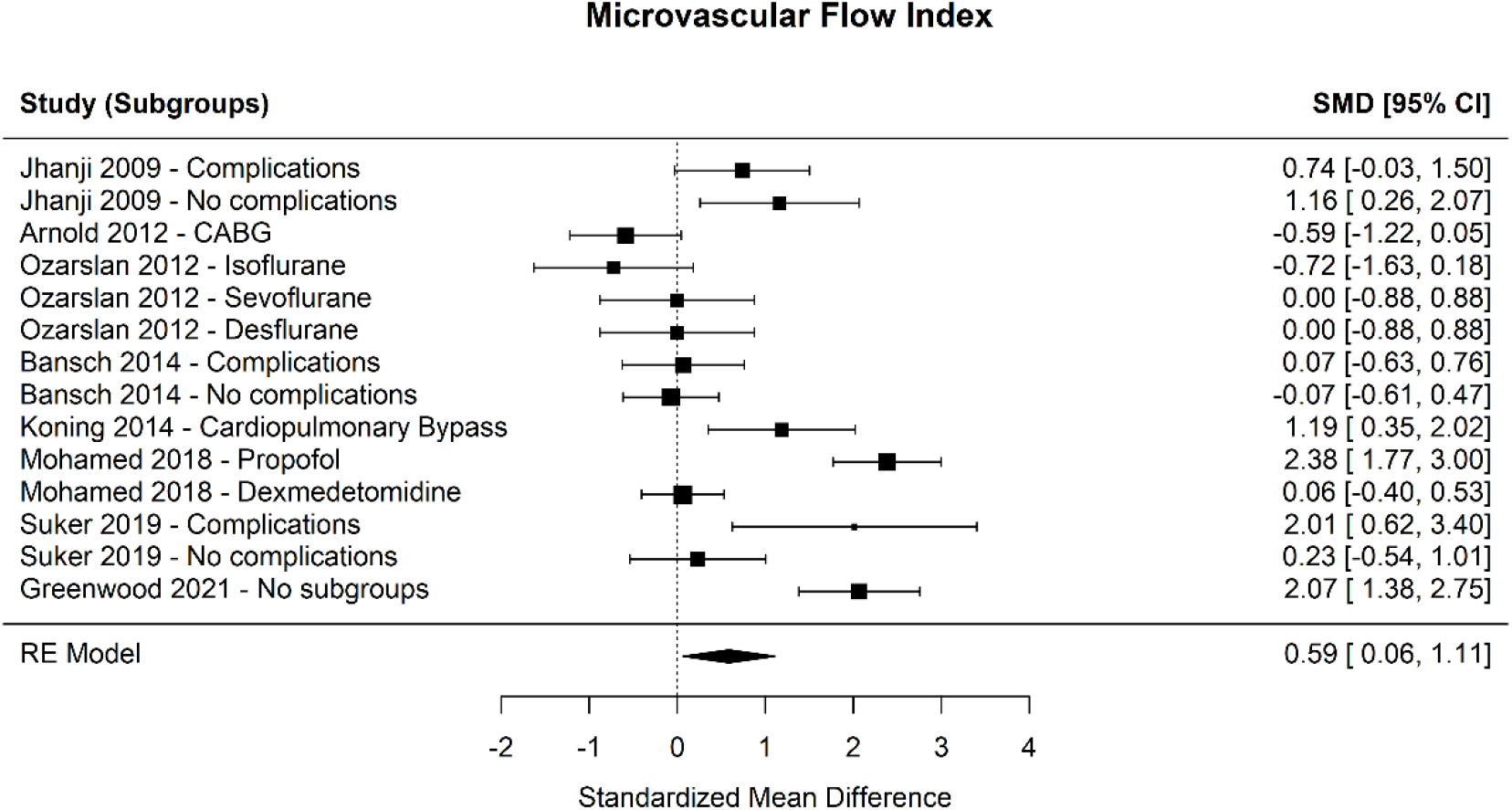
Forest plot for microvascular flow index. SMD, standardized mean difference; CI, confidence interval.

**Figure 6.**
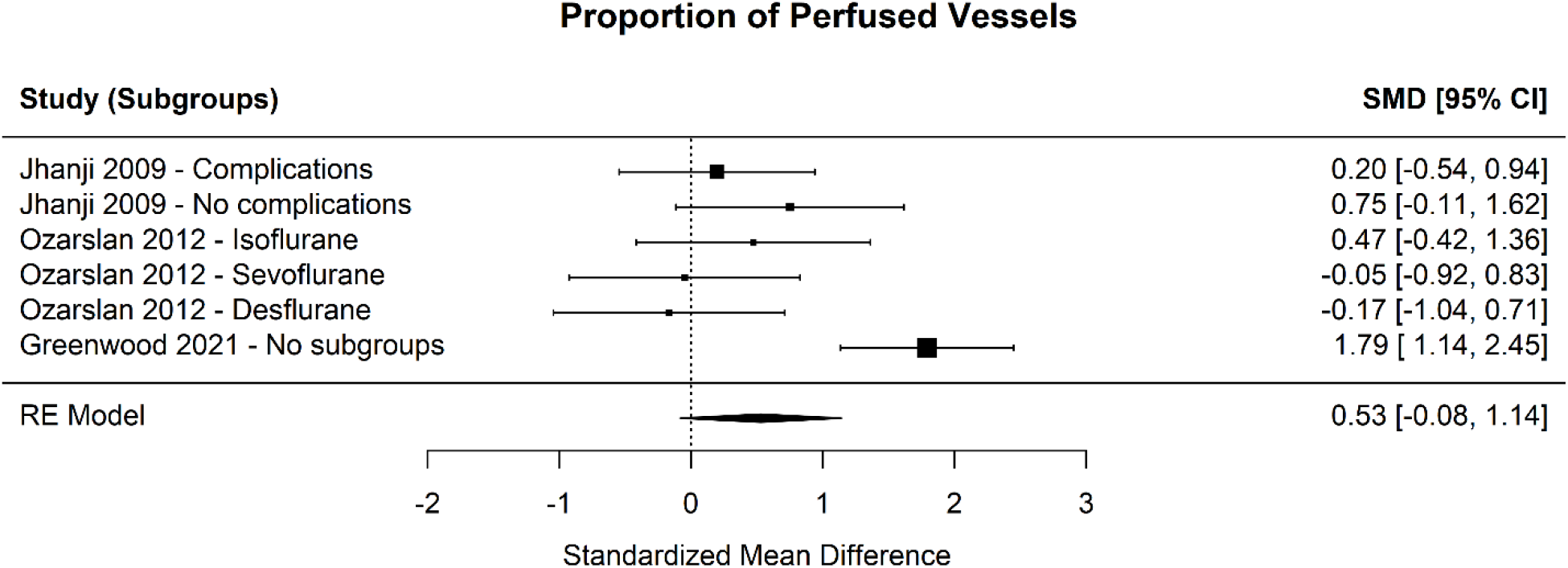
Forest plot for proportion of perfused vessels. SMD, standardized mean difference; CI, confidence interval.

### 3.2 Synthesis without cardiac surgery patients

The results in the non-cardiac surgery subgroup were comparable with the full group except that a statistically non-significant difference in PVD was found in the remaining seven studies (p=0.19; estimate: 0.26; 95% CI: −0.13 to 0.66; I^2^ = 71.94%, Q = 33.42, p=0.002) (Supplementary Table 8) [9,11-14,17]. The other parameters did not differ significantly from their respective full group results. Forest plots for the different measurements are presented in Supplementary Figures 1-5.

### 3.3 Correlation between complications and microvascular flow

As stated before, it was not possible to estimate the correlation between complications and alterations in microcirculation. The characteristics of complications in the included studies are presented in Supplementary Table 9.

### 3.4 Quality assessment and Risk of bias

Due to the nature of our analysis all studies were observational studies of exposure with no control group. There is no consensus on an established tool for the assessment of risk-of-bias in this type of studies. Most comparable scales (like the Newcastle - Ottawa) focus on two factors: a) subject and group selection, b) outcome assessment. Because this systematic review focuses on differences in the same patient, before and after exposure, group selection methods were not assessed. All included studies used randomly selected patients, but in most cases the number of study participants was low, in order to consider them a representative sample (Supplementary Table 10).

## 4. DISCUSSION

More than 310 million surgical procedures are performed annually worldwide, with the estimated postoperative mortality ranging between 1 and 4% [19,20]. Although most of the research focuses on intraoperative and postoperative hypotension, adverse cardiac events, pulmonary complications, and cognitive disorders, tissue perfusion is likely to play a key role in the development of organ dysfunction and may include systemic and/or regional alterations. Our study showed that: 1) TVD had no statistically significant difference perioperatively; 2) PVD significantly decreased postoperatively; 3) PBR significantly increased postoperatively; 4) MFI significantly decreased postoperatively; 5) PPV had no statistically significant difference perioperatively; 6) the results in the non-cardiac surgery subgroup were comparable with the full group except that a statistically non-significant difference in PVD was observed.

Comorbidities and decreased functional capacity can predispose patients to adverse surgical outcomes, while the anesthesia and perioperative stress may further aggravate the postoperative physiology. Also, the direct toxicity of pro-inflammatory mediators and/or circulating particles released from damaged cells may contribute to tissue hypoperfusion and organ dysfunction. The resulting complications are associated with increased healthcare cost and reduced long-term survival, especially in high-risk procedures. However, most physicians aim at improving the macrocirculation and ventilation parameters in an effort to maintain a balance between oxygen supply and oxygen demand, but alterations of microcirculatory perfusion may be associated with organ injury and postoperative morbidity.

Total vessel density is one of the two main functional components of microcirculation that describe its capacity to perfuse the tissues [21]. Our analysis revealed a statistically non-significant difference between preoperative and postoperative TVD, with low heterogeneity among studies. However, we found a statistically significant decrease in PVD, with medium to high heterogeneity. The method for assessing the microcirculation differed between studies, which is an important difference and a possible bias because differences in normal values for TVD between various optical systems may be up to 30% [16,21,22]. In a study of patients undergoing major abdominal surgery, those with complications experienced a decrease in PVD during the first 6 postoperative hours, which appeared to be returning to normal by 8 hours, while in those without complications, PVD returned to normal at 6 hours [8]. In a latter study by the same group, stroke volume-guided intravenous colloid therapy maintained better the microvascular flow compared to the central venous pressure-guided therapy group, while the addition of dopexamine further increased PVD [23]. Although the improvement in postoperative PVD was expected to be more evident in the dopexamine group due to its b_2_-adrenoceptor-mediated inotropic and vasodilator actions, the stroke volume-only group had the fewer complications. Perhaps other causes may be responsible for this finding. However, all groups experienced complications and this could be attributed to the increase in central venous pressure in all of them. An increased central venous pressure decreases venous return and retrogradely increases post-capillary venular pressure, which impairs capillary perfusion [24]. This could be reflected in the study by Suker et al. in which TVD was lower at 24 hours and at the 4^th^ postoperative day compared to its preoperative value, while PVD at 24 hours was practically unchanged in the complications group, but lower in the no complications group compared to its preoperative value [17]. Of note, the authors reported a fluid balance of 3983±2128 ml in the first 24 hours [17], which may have resulted in these microcirculatory alterations [21,24]. An inverse correlation between central venous pressure and microvascular flow has been reported by other authors as well [14].

Perfused boundary region describes the extent of penetration of the flowing red blood cells in μm into the luminal surface of the endothelial glycocalyx by measuring their radial motion away from the central flow towards the endothelial cells. The more the endothelial glycocalyx is injured, the deeper red blood cells penetrates into the glycocalyx and the higher the PBR is [15,25]. In our analysis, PBR was significantly increased postoperatively, without the presence of high degrees of heterogeneity. This is a very important finding because structural and functional damage to glycocalyx can be triggered in various disease states, as well as intraoperatively due to fluid overload, hypotension, transfusion, and other causes. Endothelial degradation is a cornerstone in ischemia-reperfusion-related endothelial dysfunction. It further impairs local microcirculation and disrupts normal microvascular permeability, eventually leading to acute organ injury and chronic organ failure [26-29]. Koning et al. and Astapenko et al. used sidestream dark-field imaging to assess sublingual microcirculation [11,15]. This technology has been shown to reflect the changes in the microvasculature of gastric, renal, and nervous system [30-33]. Perfused boundary region was increased postoperatively in both studies, but only Astapenko et al. reported various complications [15]. Although we could not statistically assess the aforementioned changes with the different outcomes of the study, our findings indicate that intraoperative glycocalyx injury may have important implications for patient outcomes.

Microvascular flow index is a semi-quantitative assessment of the average red blood cell velocity and expresses data on the clinical relevance of microvascular alterations. Our analysis revealed that MFI significantly decreased postoperatively, but high degrees of heterogeneity were present. Although cutoff values for a normal MFI are > 2.9, cutoff values of 2.6 are suggested as a threshold below which alterations can be considered clinically relevant [21,34,35]. As seen in supplementary table 4, most studies reached the 2.6 cutoff value. In the study by Jhanji et al., MFI value was 2.5 immediately after surgery in the complications and no complications subgroup, but remained lower than 2.6 only in the first one at 6 hours postoperatively [8]. Hospital length of stay was higher in complications group. In their later publication, MFI was ranged from 2.5 to 2.7 during the postoperative period in all subgroups, with hospital length of stay in the study being 14-16 days [23]. In a study of patients undergoing open elective pancreatico-biliary surgery, MFI was 2.98 at the 4^th^ postoperative day, but we could not compare this value with the preoperative ones or the values immediately after surgery due to data unavailability [17]. In that study, the complications group experienced a statistically non-significant decrease in MFI, while in the no complication group, the postoperative MFI value was higher than the preoperative one, but the difference was also non-significant [17]. In another study including cardiopulmonary bypass patients, perioperative MFI was constantly lower than 2.6, but none of them developed major postoperative complications [16]. Specifically, MFI at arrival to ICU was higher than the preoperative value (on the hospital floor before surgery), but MFI after transfer to the ward was lower than the ICU value (MFI_PREOP_<MFI_WARD_<MFI_ICU_) [16]. The increased MFI directly after cardiac surgery could be due to the administration of vasodilating or inotropic drugs. However, it is not clear if a baseline measurement the day before surgery could be compared with a measurement in the pre-anesthesia suite after a 6 hour or longer fasting period. Perhaps, an increase in postoperative MFI compared to the preoperative baseline value may be more important than reaching the threshold of 2.6, which has been reported to require further validation in intervention trials with clinical outcomes [21].

Our analysis revealed a statistically non-significant difference between preoperative and postoperative PPV. In a study of patients undergoing major abdominal surgery, postoperative PPV (small vessels <20 μm) was lower in those who developed complications [8]. Özarslan et al. reported transient alterations after cardiac surgery, with cardiopulmonary bypass decreasing or increasing PPV depending on the inhalational anesthetic agent [12]. Greenwood et al. also reported a decrease in PPV after elective cardiac surgery requiring cardiopulmonary bypass [14]. Indeed, cardiopulmonary bypass may induce significant changes in microvascular flow due to increased shear stress, inflammation, and fluid overload [36]. In our analysis, however, the only difference between the full group and the non-cardiac surgery group was the decrease in PVD in the first one.

## 5. RISK OF BIAS

The risk of bias in the outcome assessment rests in the accuracy of the different study methodologies. There are differences between the available technologies in optical resolution and capillary visualization (Supplementary table 9). These differences may be due to the design of the devices and suggest that measurements of microcirculatory perfusion may differ depending on the camera system used. In addition, the results of the studies are affected by the used software to analyze the videos. Therefore, the absolute measurement values may differ significantly and should always be compared very cautiously, while additional analyses and assessments are needed before a final conclusion is reached [1,3]. Today, only the second generation sidestream dark-field (SDF) imaging and the third-generation incident dark-field (IDF) imaging are available.

## 6. LIMITATIONS

This analysis investigated microvascular blood flow during the immediate and early postoperative period in cardiac and non-cardiac surgery and in various settings, i.e., the postoperative anesthesia care unit, the ICU, or the ward. Consequently, it included studies investigating heterogeneous groups of surgical patients. Due to the lack of multiple randomized controlled trials, our effort to synthesize all available knowledge on the specific outcomes was difficult. Therefore, the conclusions drawn from this review and the clinical implications must be cautious and reserved. Also, during the initial screening for this study, 14,667 articles were excluded and there is always a risk that some articles may have been falsely excluded. The inclusion criteria were based on current evidence and on what we believed that would yield the most accurate answer to our research question. In addition, as we have stated in the Data Analysis section, neither statistical assessment of publication bias (or meta-regression for the association with different study outcomes) nor subgroup analysis could be performed due to the limited number of studies included in each analysis. Finally, the analysis of PBR is a very special technique that is not yet widely available. The practical meaning of a decreased glycocalyx height in the perioperative setting has not been clarified, while the studies assessing PBR did not measure glycocalyx destruction markers in the plasma.

## 7. CONCLUSIONS

Significant sublingual microcirculatory alterations are present during the immediate and early postoperative period. Further research is required for the full clarification of their effects on postoperative complications and outcome in adult patients following surgery.

## 8. PERSPECTIVES

Microcirculatory impairment can result in inadequate tissue perfusion and organ injury. Our findings suggest that poor microvascular flow and glycocalyx injury may play an important role in the evolution of postoperative complications. Moreover, our findings support the concept that microcirculatory blood flow indices can yield important physiologic information distinct from macrocirculatory hemodynamic parameters. Further research and well-designed trials using third-generation hand-held vital microscopes are necessary in order to develop specific treatment strategies and integrate the assessment of microcirculation in perioperative care.

## Data Availability

Data can be made available upon request through a collaborative process. Please contact for additional information.

## Details of authors’ contributions

Conceptualization: AC

Formal analysis: AC, NP, GM, KK, MM, IP, EL, EA

Methodology: NP, GM, MM

Writing - original draft: AC

Writing - review and editing: AC, NP, GM, KK, MM, IP, EL, EA

All authors had full access to all of the data in the study and take responsibility for the integrity of the data and the accuracy of the data analysis.

## Acknowledgements

The authors would like to thank Dr. Nick J. Koning, Dr. Alexander B. A. Vonk, Dr. Hans Vink, and Prof. Christa Boer for responding to our request for additional study data.

## LIST OF ABBREVIATIONS

ICU: intensive care unit
IDF: incident dark-field
MFI: microvascular flow index
PBR: perfused boundary region
PPV: proportion of perfused vessels
PVD: perfused vessel density
SDF: sidestream dark-field
SMD: standardized mean differences
TVD: total vessel density

## Notes

### Competing Interest Statement

The authors have declared no competing interest.

### Funding Statement

This research did not receive any specific grant from funding agencies in the public, commercial, or not-for-profit sectors.

## REFERENCES

[1] Guven G, Hilty MP, Ince C. Microcirculation: Physiology, Pathophysiology, and Clinical Application. Blood Purif 2020;49:143–50.

[2] Ocak I, Kara A, Ince C. Monitoring microcirculation. Best Pract Res Clin Anaesthesiol 2016;30:407–18.

[3] Dilken O, Ergin B, Ince C. Assessment of sublingual microcirculation in critically ill patients: consensus and debate. Ann Transl Med 2020;8:793.

[4] Moher D, Liberati A, Tetzlaff J, Altman DG; PRISMA Group. Preferred reporting items for systematic reviews and meta-analyses: the PRISMA statement. PLoS Med 2009;6:e1000097.

[5] McGrath S, Zhao X, Steele R, Thombs BD, Benedetti A; DEPRESsion Screening Data (DEPRESSD) Collaboration. Estimating the sample mean and standard deviation from commonly reported quantiles in meta-analysis. Stat Methods Med Res. 2020; in press.

[6] Cochrane Handbook for Systematic Reviews of Intervention, https://training.cochrane.org/handbook; 2020 [Accessed 07 September 2020].

[7] De Backer D, Dubois MJ, Schmartz D, Koch M, Ducart A, Barvais L, et al. Microcirculatory alterations in cardiac surgery: effects of cardiopulmonary bypass and anesthesia. Ann Thorac Surg 2009;88:1396–403.

[8] Jhanji S, Lee C, Watson D, Hinds C, Pearse RM. Microvascular flow and tissue oxygenation after major abdominal surgery: association with post-operative complications. Intensive Care Med 2009;35:671–7.

[9] Mohamed H, Hosny H, Tawadros Md P, Elayashy Md Desa Fcai M, El-Ashmawi Md H. Effect of Dexmedetomidine Infusion on Sublingual Microcirculation in Patients Undergoing On-Pump Coronary Artery Bypass Graft Surgery: A Prospective Randomized Trial. J Cardiothorac Vasc Anesth 2019;33:334–40.

[10] O’Neil MP, Alie R, Guo LR, Myers ML, Murkin JM, Ellis CG. Microvascular Responsiveness to Pulsatile and Nonpulsatile Flow During Cardiopulmonary Bypass. Ann Thorac Surg 2018;105:1745–53.

[11] Koning NJ, Vonk AB, Vink H, Boer C. Side-by-Side Alterations in Glycocalyx Thickness and Perfused Microvascular Density During Acute Microcirculatory Alterations in Cardiac Surgery. Microcirculation 2016;23:69–74.

[12] Özarslan NG, Ayhan B, Kanbak M, Çelebioğlu B, Demircin M, Ince C, et al. Comparison of the effects of sevoflurane, isoflurane, and desflurane on microcirculation in coronary artery bypass graft surgery. J Cardiothorac Vasc Anesth 2012;26:791–8.

[13] Bansch P, Flisberg P, Bentzer P. Changes in the sublingual microcirculation during major abdominal surgery and post-operative morbidity. Acta Anaesthesiol Scand 2014;58:89–97.

[14] Greenwood JC, Jang DH, Hallisey SD, et al. Severe Impairment of Microcirculatory Perfused Vessel Density Is Associated With Postoperative Lactate and Acute Organ Injury After Cardiac Surgery. J Cardiothorac Vasc Anesth 2021;35:106–15.

[15] Astapenko D, Pouska J, Benes J, Skulec R, Lehmann C, Vink H, et al. Neuraxial anesthesia is less harmful to the endothelial glycocalyx during elective joint surgery compared to general anesthesia. Clin Hemorheol Microcirc 2019;72:11–21.

[16] Arnold RC, Dellinger RP, Parrillo JE, Chansky ME, Lotano VE, McCoy JV, et al. Discordance between microcirculatory alterations and arterial pressure in patients with hemodynamic instability. J Crit Care 2012;27:531.e1-7.

[17] Suker M, Tovar Doncel MS, Pinto Lima AA, Ince C, van Eijck CHJ. Sublingual microcirculation in pancreatico-biliary surgery: An observational study. Clin Hemorheol Microcirc 2019;72:247–57.

[18] Koning NJ, Vonk AB, Meesters MI, Oomens T, Verkaik M, Jansen EK, et al. Microcirculatory perfusion is preserved during off-pump but not on-pump cardiac surgery. J Cardiothorac Vasc Anesth 2014;28:336–41.

[19] Weiser TG, Haynes AB, Molina G, Lipsitz SR, Esquivel MM, Uribe-Leitz T, et al. Estimate of the global volume of surgery in 2012: an assessment supporting improved health outcomes. Lancet 2015;385 Suppl 2:S11.

[20] Pearse RM, Moreno RP, Bauer P, Pelosi P, Metnitz P, Spies C, et al; European Surgical Outcomes Study (EuSOS) group for the Trials groups of the European Society of Intensive Care Medicine and the European Society of Anaesthesiology. Mortality after surgery in Europe: a 7 day cohort study. Lancet 2012;380:1059–65.

[21] Ince C, Boerma EC, Cecconi M, De Backer D, Shapiro NI, Duranteau J, et al; Cardiovascular Dynamics Section of the ESICM. Second consensus on the assessment of sublingual microcirculation in critically ill patients: results from a task force of the European Society of Intensive Care Medicine. Intensive Care Med 2018;44:281–99.

[22] Vincent JL, Taccone FS. Microvascular monitoring - Do ‘global’ markers help? Best Pract Res Clin Anaesthesiol 2016;30:399–405.

[23] Jhanji S, Vivian-Smith A, Lucena-Amaro S, Watson D, Hinds CJ, Pearse RM. Haemodynamic optimisation improves tissue microvascular flow and oxygenation after major surgery: a randomised controlled trial. Crit Care 2010;14:R151.

[24] Chalkias A, Xanthos T, Papageorgiou E, Anania A, Beloukas A, Pavlopoulos F. Intraoperative initiation of a modified ARDSNet protocol increases survival of septic patients with severe acute respiratory distress syndrome. Heart Lung 2018;47:616–21.

[25] Lee DH, Dane MJ, van den Berg BM, Boels MG, van Teeffelen JW, de Mutsert R, et al; NEO study group. Deeper penetration of erythrocytes into the endothelial glycocalyx is associated with impaired microvascular perfusion. PLoS One 2014;9:e96477.

[26] Abassi Z, Armaly Z, Heyman SN. Glycocalyx Degradation in Ischemia-Reperfusion Injury. Am J Pathol 2020;190:752–67.

[27] Salmon AH, Satchell SC. Endothelial glycocalyx dysfunction in disease: albuminuria and increased microvascular permeability. J Pathol 2012;226:562–74.

[28] Brettner F, von Dossow V, Chappell D. The endothelial glycocalyx and perioperative lung injury. Curr Opin Anaesthesiol 2017;30:36–41.

[29] Giacinto O, Satriano U, Nenna A, Spadaccio C, Lusini M, Mastroianni C, et al. Inflammatory Response and Endothelial Dysfunction Following Cardiopulmonary Bypass: Pathophysiology and Pharmacological Targets. Recent Pat Inflamm Allergy Drug Discov 2019;13:158–73.

[30] Vlahu CA, Lemkes BA, Struijk DG, Koopman MG, Krediet RT, Vink H. Damage of the endothelial glycocalyx in dialysis patients. J Am Soc Nephrol 2012;23:1900–8.

[31] Martens RJ, Vink H, van Oostenbrugge RJ, Staals J. Sublingual microvascular glycocalyx dimensions in lacunar stroke patients. Cerebrovasc Dis 2013;35:451–4.

[32] Piagnerelli M, Ince C, Dubin A. Microcirculation. Crit Care Res Pract 2012;2012:867176.

[33] Verdant CL, De Backer D, Bruhn A, Clausi CM, Su F, Wang Z, et al. Evaluation of sublingual and gut mucosal microcirculation in sepsis: a quantitative analysis. Crit Care Med 2009;37:2875–81.

[34] Vellinga NA, Boerma EC, Koopmans M, Donati A, Dubin A, Shapiro NI, et al; microSOAP Study Group. International study on microcirculatory shock occurrence in acutely ill patients. Crit Care Med 2015;43:48–56.

[35] Tanaka S, Harrois A, Nicolaï C, Flores M, Hamada S, Vicaut E, et al. Qualitative real-time analysis by nurses of sublingual microcirculation in intensive care unit: the MICRONURSE study. Crit Care 2015;19:388.

[36] Moret E, Jacob MW, Ranucci M, Schramko AA. Albumin-Beyond Fluid Replacement in Cardiopulmonary Bypass Surgery: Why, How, and When? Semin Cardiothorac Vasc Anesth 2014;18:252–9.

