## Supplemental files for "Sublingual microcirculatory alterations during the immediate and early postoperative period: A systematic review and meta-analysis"

### Search strategy

### PubMed

(((("microcirculatory"[All Fields] AND (((((((("perfusable"[All Fields] OR "perfusate"[All Fields]) OR "perfusates"[All Fields]) OR "perfuse"[All Fields]) OR "perfused"[All Fields]) OR "perfuses"[All Fields]) OR "perfusing"[All Fields]) OR "perfusion"[MeSH Terms]) OR "perfusion"[All Fields]) OR "perfusions"[All Fields])) OR ("microcirculatory"[All Fields] AND "flow"[All Fields])) OR (((("microvascular"[All Fields] OR "microvascularity"[All Fields]) OR "microvascularization"[All Fields]) OR "microvascularized"[All Fields]) AND (((((((("perfusable"[All Fields] OR "perfusate"[All Fields]) OR "perfusates"[All Fields]) OR "perfuse"[All Fields]) OR "perfused"[All Fields]) OR "perfuses"[All Fields]) OR "perfusing"[All Fields]) OR "perfusion"[MeSH Terms]) OR "perfusion"[All Fields]) OR "perfusions"[All Fields]))) OR (((("microvascular"[All Fields] OR "microvascularity"[All Fields]) OR "microvascularization"[All Fields]) OR "microvascularized"[All Fields]) AND "flow"[All Fields])) AND (((((("postoperative period"[MeSH Terms] OR ("postoperative"[All Fields] AND "period"[All Fields])) OR "postoperative period"[All Fields]) OR "postop"[All Fields]) OR "postoperative"[All Fields]) OR "postoperatively"[All Fields]) OR "postoperatives"[All Fields])

### Scopus

"postoperative" AND " microcirculation"

**Supplementary Figure 1**

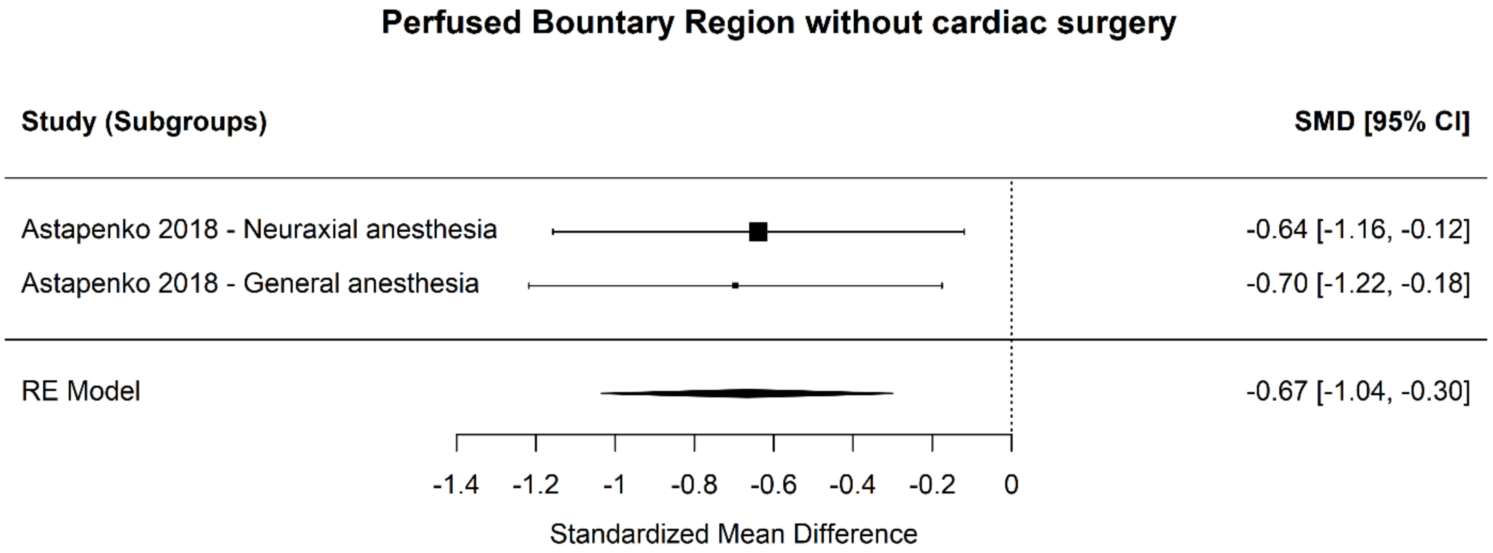

**Supplementary Figure 2**

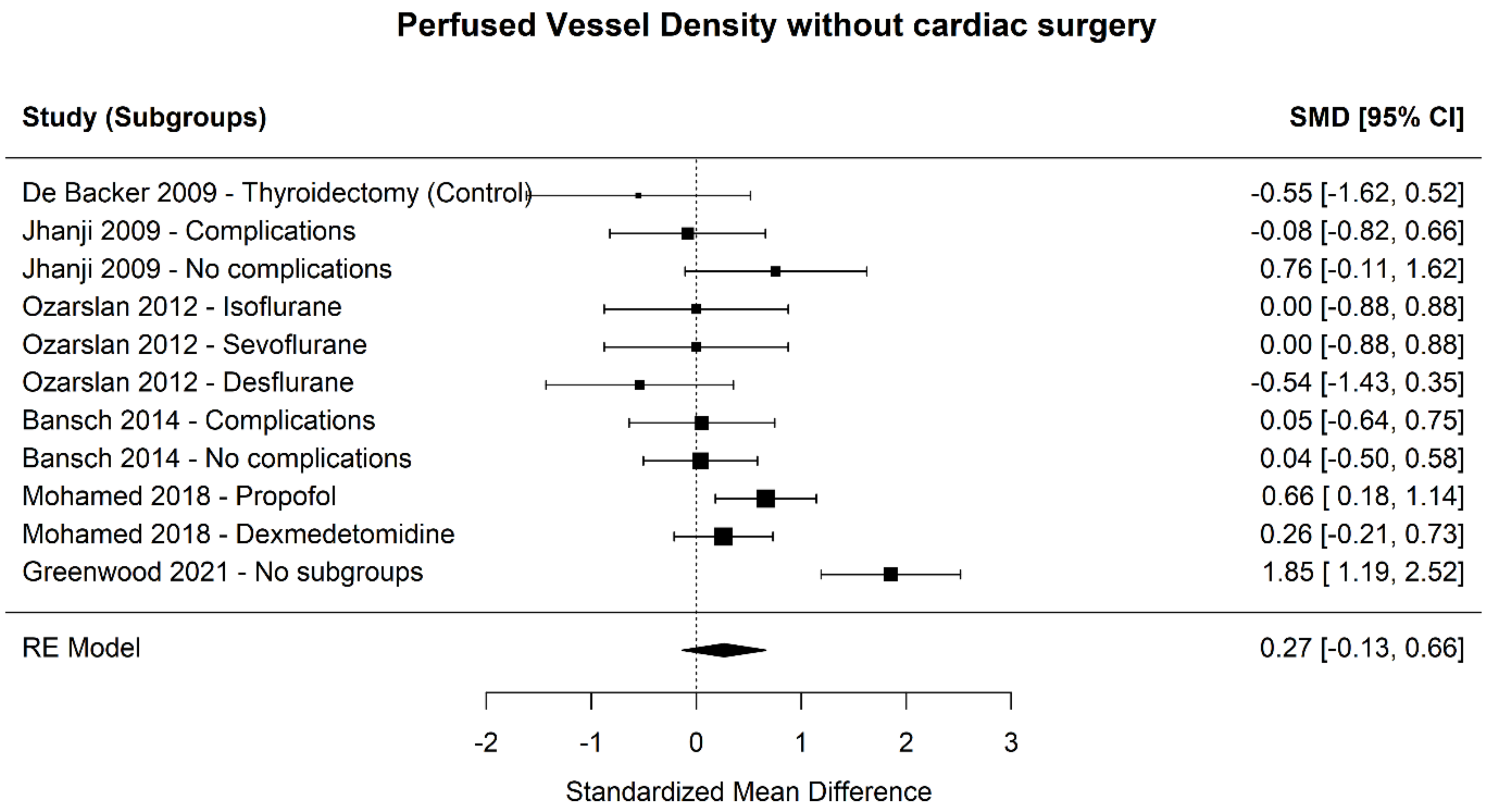

**Supplementary Figure 3**

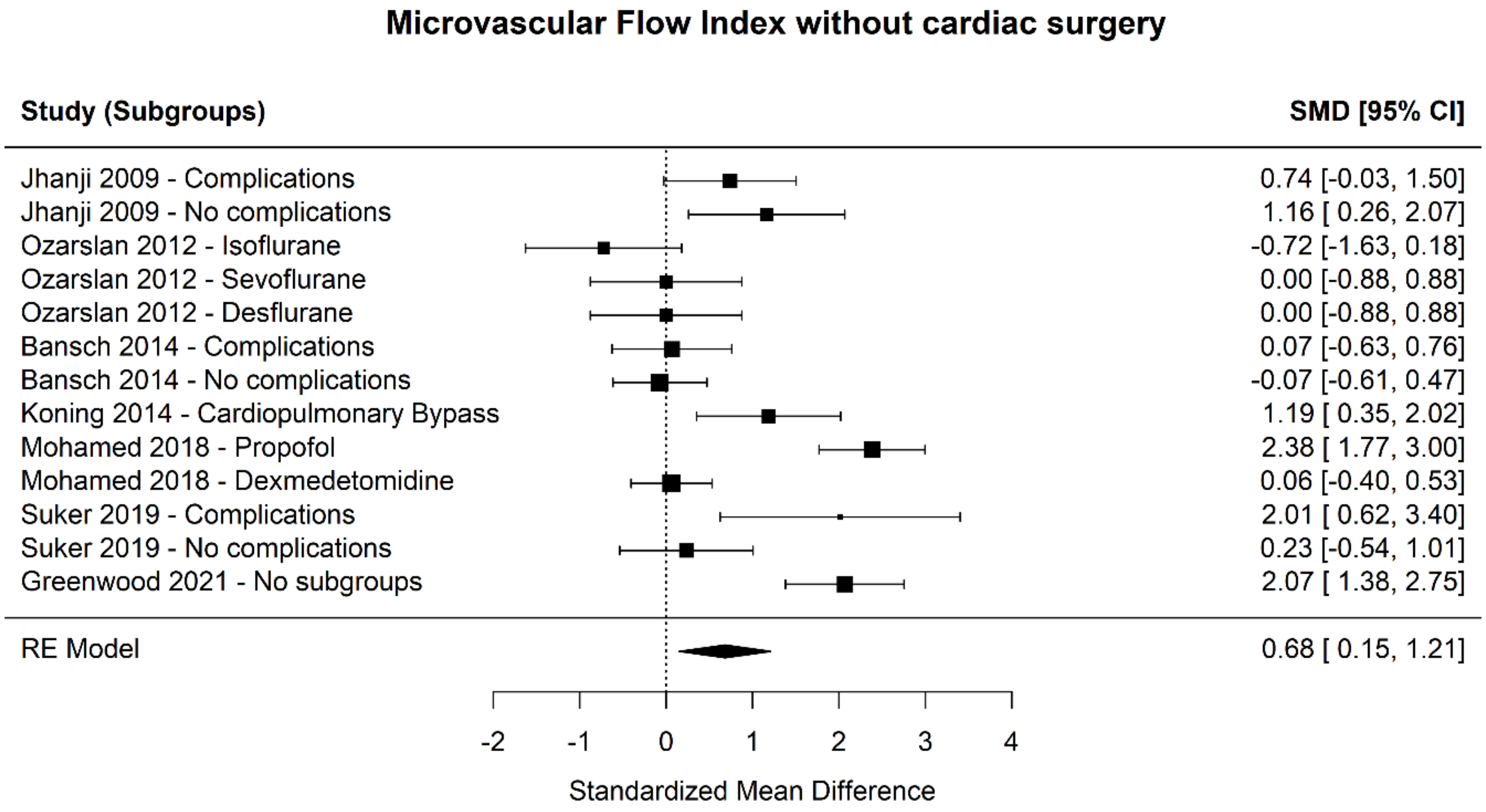

**Supplementary Figure 4**

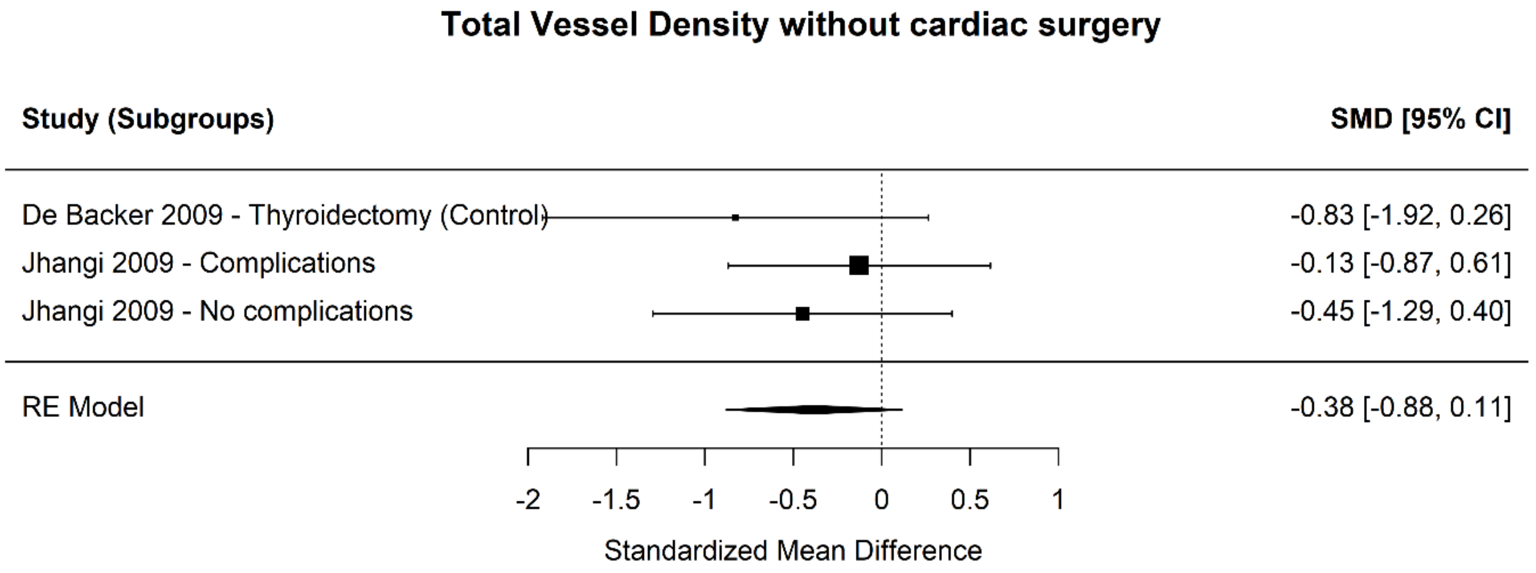

**Supplementary Figure 5**

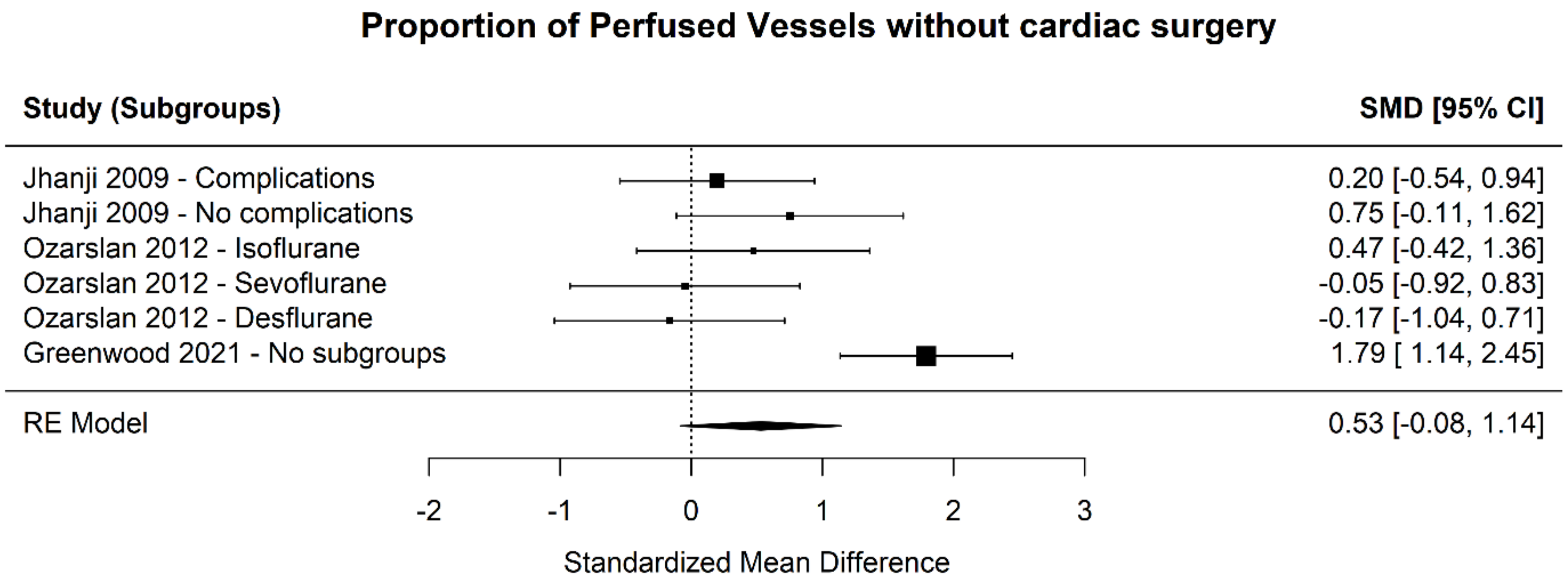

**Supplementary Table 1. Definitions of microcirculatory parameters**

| Variable | Abbreviation | Definition | Units | Characteristics |
|---|---|---|---|---|
| Total vessel density | TVD | Software supported measurement of total vessel area per surface area.<br>The proportion of vessel area with flowing blood over the total measurement area.<br>It can be calculated as the number of vessels crossing the lines divided by the total length of the lines.<br>Perfusion can then be categorized as present, absent, or intermittent. | $mm^2/mm^2$ | Determinant of capillary distance (diffusive capacity).<br>The physiological relevance of an increased vessel density is questionable as this assessment includes perfused and not perfused micro-vessels.<br>Absolute number, continuous data.<br>Exact measurements of vessel diameter. |

| | | | | |
|---|---|---|---|---|
| Perfused vessel density | PVD | Percentage of perfused vessels × total vessel density | $mm^2/mm^2$ | Determinant of capillary distance (diffusive capacity) and red blood cell velocity (convective capacity).<br>Equal to functional capillary density. |
| Perfused Boundary Region | PBR | A portion of the luminal side of the endothelial glycocalyx that is partially accessible to flowing red blood cells. | µm | It reflects the thickness of the endothelial glycocalyx.<br>Damage to glycocalyx results in increased perfused boundary region, which can be measured in human sublingual microvasculature recordings. |
| Microvascular flow index | MFI | Grid-based score per quadrant.<br>0 = stop flow, 1 = intermittent flow, 2 = sluggish flow, 3 = normal flow. | AU | Semi-quantitative assessment of the average red blood cell velocity per quadrant.<br>Good reproducibility. |

| | | | | |
|---|---|---|---|---|
| | | | | Potential loss of detail, but may overcome by similar score per vessel. |
| Proportion of perfused vessels | PPV | Grid-based score (3 horizontal and vertical equidistant lines).<br>Percentage of perfused vessels per total number of vessel crossings | % | Binominal determinant of red blood cell velocity (flow or no-flow).<br>Good reproducibility.<br>Is sensitive to isotropy. |

AU arbitrary unit

**Supplementary Table 2. Data extraction for microcirculatory parameter – Total vessel density**

| Author - year | Subgroups | N | Total vessel density | | | | | | | |
|---|---|---|---|---|---|---|---|---|---|---|
| | | | Preoperative | | Postoperative - Immediate | | Postoperative - Early | | | |
| | | | Mean | Time | Mean | Time | 1. Mean | 1. Time | 2. Mean | 2. Time |
| De Backer - 2009 | CPB | 9 | 7.5 (6.4-7.7) | Previous day | 7.6 (7.0-8.2) | Admission to ICU | 7.7 (6.7-8.3) | 6 hrs | 7.9 (6.9-8.1) | 24 hrs |
| | Off-pump | 6 | 6.8 (6.5-7.7) | | 7.7 (7.0-8.1) | | 7.4 (7.1-8.4) | | 7.8 (6.9-8.2) | |
| | Thyroidectomy (control) | 7 | 7.2 (7.0-7.7) | | 7.7 (7.6-7.8) | Admission to PACU | 7.6 (6.9-7.9) | | 7.8 (7.6-8.0) | |
| Jhangi - 2009 | Complications | 14 | 6.8 (5.5-7.8) | On the ward prior to surgery | 6.5 (5.8-7.4) | Admission to ICU | 6.3 (4.6-7.0) | 6 hrs | 6.8 (5.1-7.5) | 8 hrs |
| | No complications | 11 | 5.9 (5.7-6.5) | | 6.3 (5.3-8.2) | | 6.6 (5.9-7.6) | | 7.0 (6.5-7.5) | |
| Ozarslan - 2012 | Isoflurane | 10 | 0.6 (0.4) | Before Induction | 0.5 (0.5) | 0 hrs | 0.5 (0.5) | After 24 hrs | - | - |
| | Sevoflurane | 10 | 1.3 (0.8) | Before Induction | 1.3 (0.7) | 0 hrs | 1.4 (0.8) | After 24 hrs | - | - |
| | Desflurane | 10 | 1.0 (0.5) | Before Induction | 1.1 (1.0 | 0 hrs | 0.8 (0.5) | After 24 hrs | - | - |
| Mohamed - 2018 | Propofol | 35 | 7.6 (6.3-9.9) | Before bypass | 7.24 (1.66) | 30 min after weaning | - | - | - | - |
| | Dexmedetomidine | 35 | 10.1 (8.8-13.9) | Before bypass | 9.27 (2.27) | 30 min after weaning | - | - | - | - |
| O’Neil - 2018 | Pulsatile | 10 | 13.18 (0.67) | Before Induction | 12.79 (0.41) | 1 hrs after surgery | 13.46 (0.56) | After 24 hrs | - | - |

| | | | | | | | | | | |
|---|---|---|---|---|---|---|---|---|---|---|
| | Non pulsatile | 10 | 12.47 (0.51) | Before Induction | 12.67 (0.41) | 1 hrs after surgery | 13.79 (0.48) | After 24 hrs | | |
| Suker - 2019 | Clavien-Dindo III or higher complications: intra-abdominal abscess: 3; pancreatic fistula: 1; bile leakage: 1; liver necrosis: 1 | 6 | 26 (5.2) | 1 day before surgery | Not assessed | Not assessed | 20.7 (3.2) | After 24 hrs | 23.52 (1.42) | 4th postop day |
| | No complications | 13 | 24 (5) | | Not assessed | | 22.8 (6.3) | After 24 hrs | | |

CPB, cardiopulmonary bypass; ICU, intensive care unit; PACU, post-anesthesia care unit

**Supplementary Table 3. Data extraction for microcirculatory parameter – Perfused vessel density**

| Author - year | Type of study | Population | Microcirculatory assessment | Subgroups | M/F | Age | N | Perfused vessel density | | | | | | | | Hospital length of stay | Postop complications | Comments |
|---|---|---|---|---|---|---|---|---|---|---|---|---|---|---|---|---|---|---|
| | | | | | | | | Preoperative | | Postoperative - Immediate | | Postoperative - Early | | | | | | |
| | | | | | | | | Mean | Time | Mean | Time | 1. Mean | 1. Mean | 2. Mean | 2. Time | | | |
| De Backer - 2009[8] | Prospective | Cardiac surgery | Cytoscan A/RII - Five sequences of 20 seconds each | CPB | 6/3 | Median: 66 | 9 | Median: 4.3 (3.8-5.1) | 1 day before surgery | 3.1 (2.8-3.6) | Admission to ICU | 3.8 (2.8-4.4) | 6 hrs | 4.4 (3.9-4.7) | 24 hrs | Not assessed | Only the SOFA score is reported | |
| | | | | Off-pump | 6/0 | Median: 66 | 6 | Median: 4.6 (4.4-5.1) | | 3.7 (3.2-4.1) | | 4.0 (3.8-4.7) | | 4.8 (3.9-5.1) | | Not assessed | | |
| | | Thyreoidectomy | | Thyroidectomy (Control) | 3/4 | Median: 59 | 7 | Median: 4.5 (4.4-4.9) | | 5.0 (4.8-5.1) | Admission to PACU | 5.0 (4.3-5.3) | | 5.1 (4.9-5.4) | | Not assessed | | |
| Jhanji - 2009[9] | Prospective | Major abdominal | Sidestream dark-field imaging with a 5x objective lens (Microscan) | Complications: 10 infectious, 2 gastrointestinal tract, 1 cardiovascular, 1 respiratory | 6/8 | Median: 69 | 14 | Median: 5.7 (4.1-6.9) | On the ward prior to surgery | Median: 4.5 (3.8-6.2) | Admission to ICU | Median: 4.1 (3.1-6.2) | 6 hrs | Median: 6.3 (3.7-6.8) | 8 hrs | Median: 20 (14-26) | Infectious: 10; gastrointestinal tract: 2; cardiovascular: 1, respiratory: 1 | |
| | | | | No complications | 5/6 | Median: 66 | 11 | Median: 5.6 (4.8-5.8) | | Median: 4.9 (3.6-6.3) | | Median: 5.7 (5.1-7.2) | | Median: 6.6 (5.2-7.3) | | Median: 8 (8-9.5) | | |
| | | | | CVP: Colloids administered to achieve rise in CVP of at least 2 mmHg for 20 minutes or more | 30/15 | Median: 70 | 45 | NA | | 6.1 (2.4) | | 5.8 (1.9) | | 5.3 (1.8) | | Median: 15 (10-26) | (Within 7 days) Cardiovascular: 4; infectious: 29; acute kidney injury: 10; others: 10 | |
| Jhanji - 2010[18] | Prospective | Major abdominal | Sidestream dark-field imaging with a ×5 objective lens (Microscan) | SV: Colloids to achieve an optimal value of stroke volume | 31/14 | Median: 68 | 45 | NA | Before surgery | 5.8 (2.5) | 0 hrs | 5.7 (1.9) | 6 hrs | 6.2 (3.0) | 8 hrs | Median: 14 (11-26) | (Within 7 days) Cardiovascular: 3; infectious: 24; acute kidney injury: 13; others: 14 | Microvessels < 20 µm |
| | | | | SV + Dopexamine | 28/27 | Median: 65 | 45 | NA | | 5.8 (2.4) | | 6.2 (1.8) | | 6.3 (3.0) | | Median: 16 (11-28) | (Within 7 days) Cardiovascular: 3; infectious: 28; acute kidney | |

| | | | | | | | | | | | | | | | | | | |
|---|---|---|---|---|---|---|---|---|---|---|---|---|---|---|---|---|---|---|
| | | | | | | | | | | | | | | | | | injury: 4; others: 12 | |
| | | | | Isoflurane | 8/2 | Mean: 59.6 (11.2) | 10 | 0.3 (0.2) | Before induction | 0.3 (0.2) | 0 hrs | 0.3 (0.2) | After 24 hrs | - | - | - | - | - |
| Ozarslan - 2012 | RCT | CABG | Orthogonal polarization spectral imaging sublingual | Sevoflurane | 9/1 | Mean: 67.7 (4.6) | 10 | 0.4 (0.4) | Before induction | 0.4 (0.3) | 0 hrs | 0.4 (0.4) | After 24 hrs | - | - | - | - | - |
| | | | | Desflurane | 8/2 | Mean: 61 (10.1) | 10 | 0.4 (0.1) | Before induction | 0.8 (1.0) | 0hrs | 0.4 (0.2) | After 24 hrs | - | - | - | - | - |
| Bansch - 2014 | Prospective | Major Abdominal Surgery | Sidestream dark field imaging sublingual | Complications | 7/9 | Median: 66 (43-86) | 16 | 12.6 (11.4-15.1) | Before induction | 12.4 (10.2-15.2) | 2 hrs after arrival at recovery room | 12.7 (8.8-16.6) | Morning after surgery | - | - | - | Infections, Respiratory, cardiovascular, GI, renal | - |
| | | | | No complications | 17/11 | Median: 64 (43-86) | 26 | 12.7 (9.7-14.9) | Before induction | 12.4 (9.7-14.6) | 2 hrs after arrival at recovery room | 12.5 (10.0-14.8) | Morning after surgery | - | - | - | - | - |
| | | | | Non-pulsatile CPB | | Median: 65 | 12 | 19.8 (2.8) | | 15.3 (2.6) | | | | | | | | |
| Koning - 2016[10] | Prospective | Cardiac surgery | Sidestream dark-field imaging | Pulsatile CPB | NA | Median: 74 | 12 | 20.86 (2.39) | After anesthesia induction | 20.3 (2.31) | After closure of sternal wound - Admission to ICU | Not assessed | Not assessed | Not assessed | Not assessed | Not assessed | Not assessed | Microvessels 5 - 25 µm |
| | | | | Off-pump | | Median: 65 | 12 | 19.8 (2.8) | | 19.4 (2.3) | | | | | | | | |
| Mohamed - 2018 | Prospective | CABG | Sidestream dark field imaging sublingual | Propofol | 29/6 | Mean: 59 (4) | 35 | 4.3 (2.7-6.2) median | Before Bypass | 3.3 (2.2-4.3) | 30 min after weaning | - | - | - | - | - | - | |
| | | | | Dexmedetomidine | 23/12 | Mean: 58 (7) | 35 | 5.7 (2.2-13.8) median | Before bypass | 6.1 (3-8.9) | 30 min after weaning | - | - | - | - | - | - | |
| Dekker - 2019[7]* | Prospective | Cardiac surgery | Sidestream dark-field imaging | CABG with CPB | 15/2 | Median: 69 | 17 | NA | NA | NA | NA | NA | NA | NA | NA | Median: 6 (5-8) | Not assessed | Microvessels 5 - 25 µm |
| Suker - 2019[13] | Prospective | Pancreatic surgery (Whipple, distal | Incident dark-field (IDF) | Clavien-Dindo III or higher complications: intra-abdominal abscess: | 11/8 | Median: 58.3 | 6 | 20.3 (4.7) | 1 day before surgery | Not assessed | Not assessed | 20.2 (6.9) | At 24 hrs | 21.14 (1.40) | $4^{th}$ postop day | Not assessed | Clavien-Dindo III or higher complications: intra-abdominal | Microvessels < 20 µm |

| | | | | | | | | | | | | | | | | | | |
|---|---|---|---|---|---|---|---|---|---|---|---|---|---|---|---|---|---|---|
| | | pancreatectomy, adhesiolysis) | imaging (Cytocam) | 3; pancreatic fistula: 1; bile leakage: 1; liver necrosis: 1 | | | | | | | | | | | | | abscess: 3; pancreatic fistula: 1; bile leakage: 1; liver necrosis: 1 | |
| | | | | No complications | | | 13 | 25.1 (5.5) | | Not assessed | Not assessed | 16.9 (3.5) | | | | Not assessed | | |
| Greenwood - 2021 | Prospective | CPB | Sidestream dark field imaging sublingual | No subgroups | 20/5 | Mean: 63 (12) | 25 | 20.7 (19.3-22.9) | Before surgery | 16.3 (12.8-17.9) | ICU, 2 hrs after surgery | - | - | - | - | - | - | - |

CPB, cardiopulmonary bypass; ICU, intensive care unit; PACU, post-anesthesia care unit; NA, data non-available; CVP, central venous pressure; SV, stroke volume; CABG: Cardiopulmonary Bypass with graft

* Parameters were reported, but only in charts (exact numbers not available)

**Supplementary Table 4. Data extraction for microcirculatory parameter – Perfused boundary region**

| | | | | | | | | Perfused boundary region | | | | | | | | | | |
|---|---|---|---|---|---|---|---|---|---|---|---|---|---|---|---|---|---|---|
| Author - year | Type of study | Population | Microcirculatory assessment | Subgroups | M/F | Age | N | Preoperative | | Postoperative - Immediate | | Postoperative - Early | | | | Postop complications | Hospital length of stay | Comments |
| | | | | | | | | Mean | Time | Mean | Time | 1. Mean | 1. Time | 2. Mean | 2. Time | | | |
| Koning - 2016[10] | Prospective | Cardiac surgery | Imaging-based digital analysis system (GlycocheckTM glycocalyx measurement software) | Nonpulsatile CPB | NA | Median: 65 | 12 | 2.4 (0.35) | After anesthesia induction | 2.6 (0.31) | After closure of sternal wound - Admission to ICU | Not assessed | Not assessed | Not assessed | Not assessed | | Not assessed | Microvessels 5 - 25 µm |
| | | | | Pulsatile CPB | | Median: 74 | 12 | 2.41 (0.26) | | 2.47 (0.19) | | | | | | | | |
| | | | | Off-pump | | Median: 65 | 12 | 2.46 (0.4) | | 2.3 (0.25) | | | | | | | | |
| Astapenko - 2019[11] | Prospective | Elective total knee/hip replacement | Microcirculation videos by specialized hand-held video microscope (GlycoCheck, Maastricht, the Netherlands). | Neuraxial anesthesia | 16/14 | 67.13 (9.81) | 30 | 1.95 (0.24) | Before surgery | 2.09 (0.19) | 2 hrs | Not assessed | Not assessed | Not assessed | Not assessed | Pulmonary embolism: 1, Myocardial infarction:1 | Not assessed | Microvessels 5 - 25 µm |
| | | | | General anesthesia | 14/16 | 66.63 (7.08) | 30 | 2.02 (0.26) | | 2.2 (0.25) | | Not assessed | Not assessed | Not assessed | Not assessed | Deep venous thrombosis:1, Supraventricular tachycardia:1 | Not assessed | |
| Dekker - 2019[7] * | Prospective | Cardiac surgery | Sidestream dark-field imaging (GlycocheckTM glycocalyx measurement software) | CABG with CPB | 15/2 | Median: 69 | 17 | NA | NA | NA | NA | NA | NA | NA | NA | | Median: 6 (5-8) | Microvessels 5 - 25 µm |

NM, non-mentioned; CPB, cardiopulmonary bypass; CABG, coronary artery bypass graft; NA, data non-available

* Parameters were reported, but only in charts (exact numbers not available)

**Supplementary Table 5. Data extraction for microcirculatory parameter – Microvascular flow index**

| Author - year | Type of study | Population | Microcirculatory assessment | Subgroups | M/F | Age | N | Microvascular flow index | | | | | | | | Postop complications | Hospital length of stay | Comments |
|---|---|---|---|---|---|---|---|---|---|---|---|---|---|---|---|---|---|---|
| | | | | | | | | Preoperative | | Postoperative - Immediate | | Postoperative - Early | | | | | | |
| | | | | | | | | Mean | Time | Mean | Time | 1. Mean | 1. Time | 2. Mean | 2. Time | | | |
| Jhanji - 2009[9] | Prospective | Mixed (major abdominal) | Sidestream darkfield imaging with a 5x objective lens (Microscan) | Complications | 6/8 | Median: 69 | 14 | Median: 2.6 (2.5-2.9) | On the ward prior to surgery | Median: 2.5 (1.9-2.8) | Admission to ICU | Median: 2.3 (2.0-2.7) | 6 hrs | Median: 2.7 (1.9-2.9) | 8 hrs | Infectious: 10; gastrointestinal tract: 2; cardiovascular: 1, respiratory: 1 | Median: 20 (14-26) | |
| | | | | No complications | 5/6 | Median: 66 | 11 | Median: 3.0 (3.0-3.0) | | Median: 2.5 (2.4-2.8) | | Median: 2.8 (2.6-3.0) | | Median: 2.6 (2.3-2.9) | | | Median: 8 (8-9.5) | |
| Jhanji - 2010[18] | Prospective | Major abdominal | Sidestream darkfield imaging with a ×5 objective lens (Microscan) | CVP: Colloids administered to achieve rise in CVP of at least 2 mmHg for 20 minutes or more | 30/15 | Median: 70 | 45 | NA | Before surgery | 2.5 (0.3) | 0 hrs | 2.6 (0.4) | 6 hrs | 2.5 (0.5) | 8 hrs | (Within 7 days) Cardiovascular: 4; infectious: 29; acute kidney injury: 10; others: 10 | Median: 15 (10-26) | |
| | | | | SV: Colloids to achieve an optimal value of stroke volume | 31/14 | Median: 68 | 45 | NA | | 2.5 (0.4) | | 2.7 (0.3) | | 2.6 (0.4) | | (Within 7 days) Cardiovascular: 3; infectious: 24; acute kidney injury: 13; others: 14 | Median: 14 (11-26) | |

| | | | | | | | | | | | | | | | | | | |
|---|---|---|---|---|---|---|---|---|---|---|---|---|---|---|---|---|---|---|
| | | | | SV + Dopexamine | 28/27 | Median: 65 | 45 | NA | | 2.5 (0.4) | | 2.7 (0.3) | | 2.5 (0.4) | | (Within 7 days) Cardiovascular: 3; infectious: 28; acute kidney injury: 4; others: 12 | Median: 16 (11-28) | |
| Arnold - 2012[12] | Prospective | Cardiac surgery | Sidestream darkfield videomicroscopy (Microscan) | CABG, Valve replacement/repair | 18/2 | 64 (6) | 20 | 2.16 (0.29) | On the hospital floor before surgery | 2.45 (0.62) | Arrival to ICU | 2.26 (0.25) | After transfer from ICU to ward | Not assessed | Not assessed | None developed major postop complications | Not assessed | |
| Ozarslan - 2012 | RCT | CABG | Orthogonal polarization spectral imaging sublignual | Isoflurane | 8/2 | Mean: 59.6 (11.2) | 10 | 3.0 (0.3) | Before induction | 3.25 (0.36) | 0 hrs | 3.0 (0.3) | After 24 hrs | - | - | - | - | |
| | | | | Sevoflurane | 9/1 | Mean: 67.7 (4.6) | 10 | 3.0 (0.11) | Before induction | 3.0 (0.21) | 0 hrs | 3.0 (0.2) | After 24 hrs | - | - | - | - | |
| | | | | Desflurane | 8/2 | Mean: 61 (10.1) | 10 | 3.5 (0.42) | Before induction | 3.5 (0.36) | 0 hrs | 3.4 (0.4) | After 24 hrs | - | - | - | - | |
| Bansch - 2014 | Prospective | Major Abdominal Surgery | Sidestream dark field imaging sublingual | Complications | 7/9 | Median: 66 (43-86) | 16 | 2.7 (2.1-3.0 | Before induction | 2.7 (1.9-3.0) | 2 hrs after arrival at recovery room | 2.7 (2.0-3.0) | Morning after surgery | - | - | Infections, Respiratory, cardiovascular, GI, renal | - | |
| | | | | No complications | 17/11 | Median: 64 (43-86) | 26 | 2.7 (2.0-3.0) | Before induction | 2.7 (2.1-3.0) | 2 hrs after arrival at recovery room | 2.7 (2.0-3.0) | Morning after surgery | - | - | - | - | |
| Koning - 2014 | Prospective | Cardiopulmonary bypass for CABG | Sidestream dark field imaging sublingual | Cardiopulmonary Bypass | 12/1 | Median: 65 (62-72) | 13 | 2.8 (2.7-2.9) | After arterial line | 2.4 (2.3-2.7) | ICU admission | - | - | - | - | - | - | Raw data only for MFI |
| Mohamed - 2018 | Prospective | CABG | Sidestream dark field imaging sublingual | Propofol | 29/6 | Mean: 59 (4) | 35 | 2.35 (0.42) | Before bypass | 1.47 (0.3) | 30 min after weaning | - | - | - | - | - | - | |

| | | | | | | | | | | | | | | | | | | |
|---|---|---|---|---|---|---|---|---|---|---|---|---|---|---|---|---|---|---|
| | | | | Dexmedetomidine | 23/12 | Mean: 58 (7) | 35 | 2.22 (0.33) | Before bypass | 2.2 (0.29) | 30 mean after weaning | - | - | - | - | - | - | |
| Suker - 2019[13] | Prospective | Pancreatic surgery (Whipple, distal pancreatectomy, adhesiolysis) | Incident dark-field (IDF) imaging (Cytocam) | Clavien-Dindo III or higher complications: intra-abdominal abscess: 3; pancreatic fistula: 1; bile leakage: 1; liver necrosis: 1 | 11/8 | Median: 58.3 | 6 | 3.0 (3.0-3.0) * | 1 day before surgery | Not assessed | Not assessed | 2.9 (2.5-3.0) * | At 24 hrs | 2.98 (0.03) | 4th postop day | Clavien-Dindo III or higher complications: intra-abdominal abscess: 3; pancreatic fistula: 1; bile leakage: 1; liver necrosis: 1 | Not assessed | Microvessels <20 µm |
| | | | | No complications | | | 13 | 2.8 (2.6-3.0) * | | Not assessed | Not assessed | 3.0 (2.5-3.0) * | | | | | | |
| Greenwood - 2021 | Prospective | CPB | Sidestream dark field imaging sublingual | no subgroups | 20/5 | Mean: 63 (12) | 25 | 2.9 (2.8-2.9) | Before surgery | 2.5 (2.4-2.7 | ICU (2 hrs after surgery) | - | - | - | - | - | - | |

CVP, central venous pressure; NA, data non-available; SV, stroke volume; CABG, coronary artery bypass graft; ICU, intensive care unit; CPB: Cardiopulmonary Bypass

* Median (25th-75th percentile)

**Supplementary Table 6. Data extraction for microcirculatory parameter – Proportion of perfused vessels**

| Author - year | Type of study | Population | Microcirculatory assessment | Type of surgical procedure/Subgroups | M/F | Age | N | Proportion of perfused vessels | | | | | | | | Hospital length of stay | Comments |
|---|---|---|---|---|---|---|---|---|---|---|---|---|---|---|---|---|---|
| | | | | | | | | Preoperative | | Postoperative - Immediate | | Postoperative - Early | | | | | |
| | | | | | | | | Mean | Time | Mean | Time | 1. Mean | 1. Time | 2. Mean | 2. Time | | |
| Jhanji - 2009[9] | Prospective | Mixed (major abdominal) | Sidestream dark-field imaging with a 5x objective lens (Microscan) | Complications | 6/8 | Median: 69 | 14 | Median: 79 (73-92) | In the ward, prior to surgery | Median: 74 (61-90) | Admission to ICU | Median: 75 (60-83) | 6 hrs | Median: 87 (78-100) | 8 hrs | Median: 20 (14-26) | |
| | | | | No complications | 5/6 | Median: 66 | 11 | Median: 89 (83-95) | | Median: 80 (61-93) | | Median: 90 (78-97) | | Median: 95 (83-97) | | Median: 8 (8-9.5) | |
| De Backer - 2009[8] | Prospective | Cardiac surgery | Cytoscan A/RII - Five sequences of 20 seconds each | CPB | 6/3 | Median: 66 | 9 | NA | 1 day before surgery | NA | Admission to ICU | NA | 6hrs | NA | 24 hrs | Not assessed | |
| | | | | Off-pump | 6/0 | Median: 66 | 6 | NA | | NA | | NA | | NA | | Not assessed | |
| | | Thyreoidectomy | | Thyreoidectomy (control) | 3/4 | Median: 59 | 7 | NA | | NA | Admission to PACU | NA | | NA | | Not assessed | |
| Jhanji - 2010[18] | Prospective | Major abdominal | Sidestream dark-field imaging with a 5x objective lens (Microscan) | CVP: Colloids administered to achieve rise in CVP of at least 2 mmHg for 20 minutes or more | 30/15 | Median: 70 | 45 | NA | Before surgery | 83 (14) | 0 hrs | 82 (18) | 6 hrs | 81 (18) | 8 hrs | Median: 15 (10-26) | Microvessels < 20 μm |

| | | | | | | | | | | | | | | | | | |
|---|---|---|---|---|---|---|---|---|---|---|---|---|---|---|---|---|---|
| | | | | SV: Colloids to achieve an optimal value of stroke volume | 31/14 | Median: 68 | 45 | NA | | 80 (15) | | 84 (13) | | 80 (19) | | Median: 14 (11-26) | |
| | | | | SV + Dopexamine | 28/27 | Median: 65 | 45 | NA | | 81 (16) | | 85 (12) | | 87 (17) | | Median: 16 (11-28) | |
| | | | | Isoflurane | 8/2 | Mean: 59.6 (11.2) | 10 | 4.6 (4.4) | Before Induction | 2.7 (3.2) | 0 hrs | 2.8 (2.4) | After 24 hrs | - | - | - | |
| Ozarslan - 2012 | RCT | CPB | Orthogonal polarization spectral imaging sublignual | Sevoflurane | 9/1 | Mean: 67.7 (4.6) | 10 | 9.4 (5.7) | Before Induction | 9.7 (6.2) | 0 hrs | 9.7 (6.0) | After 24 hrs | - | - | - | |
| | | | | Desflurane | 8/2 | Mean: 61 (10.1) | 10 | 5.0 (2.6) | Before Induction | 5.7 (5.1) | 0 hrs | 4.4 (3.1) | After 24 hrs | - | - | - | |
| Suker - 2019[13] | Prospective | Pancreatic surgery (Whipple, distal pancreatectomy, adhesiolysis) | Incident dark-field (IDF) imaging (Cytocam) | Clavien-Dindo III or higher complications: intra-abdominal abscess: 3; pancreatic fistula: 1; bile leakage: 1; liver necrosis: 1 | 11/8 | Median: 58.3 | 6 | NA | 1 day before surgery | Not assessed | Not assessed | NA | Within 24 hrs | 2.98 (0.03) | 4$^{th}$ postop day | Not assessed | Microvessels < 20 µm |
| | | | | No complications | | | 13 | NA | | Not assessed | Not assessed | NA | | | | Not assessed | |
| Dekker - 2019[7*] | Prospective | Cardiac surgery | Sidestream dark-field imaging | CABG with CPB | 15/2 | Median: 69 | 17 | NA | NA | NA | NA | NA | NA | NA | NA | Median: 6 (5-8) | |

| | | | | | | | | | | | | | | | | | |
|---|---|---|---|---|---|---|---|---|---|---|---|---|---|---|---|---|---|
| Greenwood - 2021 | Prospective | CPB | Sidestream dark field imaging sublingual | No subgroups | 20/5 | Mean: 63 (12) | 25 | 94.3 (92-96.3) | Before surgery | 76.9 (68.5-85.0) | ICU, 2 hrs after surgery | - | - | - | - | - | - |

ICU, intensive care unit; CPB, cardiopulmonary bypass; NA, data non-available; PACU, post-anesthesia care unit; CVP, central venous pressure; SV, stroke volume

* Parameters were reported, but only in charts (exact numbers not available)

**Supplementary Table 7. Summary of meta-analysis results**

| Parameter | Number of subgroups | N total | Estimate (log SMD) | p-value | 95% CI | $I^2$ | Q | p (Q) |
|---|---|---|---|---|---|---|---|---|
| TVD | 4 (9) | 137 | -0.029 | 0.844 | -0.31 to 0.26 | 22.55% | 10.23 | 0.24 |
| PVD | 6 (16) | 250 | 0.344 | 0.035* | 0.02 to 0.66 | 65.66% | 41.77 | <0.001 |
| PBR | 2 (5) | 96 | -0.415 | 0.031* | -0.79 to -0.03 | 37.21% | 6.56 | 0.16 |
| MFI | 8 (14) | 284 | 0.587 | 0.028* | 0.06 to 1.11 | 86.09% | 96.28 | <0.001 |
| PPV | 3 (6) | 80 | 0.53 | 0.089 | -0.08 to 1.14 | 70.71% | 18.99 | 0.002 |

TVD, total vessel density; PVD, perfused vessel density; PBR, perfused boundary region; MFI, microvascular flow index; PPV, proportion of perfused vessels.

**Supplementary Table 8. Summary of meta-analysis results, excluding studies involving cardiac surgery**

| Parameter | Number of subgroups | N total | Estimate (log SMD) | p-value | 95% CI | $I^2$ | Q | p (Q) |
|---|---|---|---|---|---|---|---|---|
| TVD | 2 (3) | 32 | -0.382 | 0.132 | -0.88 to 0.11 | 0% | 1.11 | 0.57 |
| PVD | 6 (11) | 199 | 0.19 | 0.266 | -0.13 to 0.66 | 71.94% | 33.42 | 0.002 |
| PBR | 1 (2) | 60 | -0.667 | <0.001*** | -1.04 to -0.30 | 0% | 0.024 | 0.88 |
| MFI | 7 (13) | 224 | 0.682 | 0.0115* | 0.15 to 1.21 | 85.04% | 83.35 | <0.001 |
| PPV | 3 (6) | 80 | 0.53 | 0.089 | -0.08 to 1.14 | 70.71% | 18.99 | 0.002 |

TVD, total vessel density; PVD, perfused vessel density; PBR, perfused boundary region; MFI, microvascular flow index; PPV, proportion of perfused vessels.

**Supplementary Table 9. Characteristics of complications**

| Study | Type of surgery | N | Mortality n (%) | Hospital length of stay (days), median (IQR) | Patients with complications n (%) | Complications, number of patients (%) |
|---|---|---|---|---|---|---|
| Jhanji - 2009[8] | Major abdominal | 25 | 2 (8) | 20 (14-26) | 14 (56) | Infectious 10 (40), Gastrointestinal 2 (8) Cardiovascular 1 (4), Respiratory 1 (4), Neurological 1 (4) |
| Jhanji - 2010[17]- CVP subgroup | Major abdominal | 45 | 6 (13) | 15 (10-26) | 30 (67) | Cardiovascular 4 (9), Infectious 29 (64), Acute kidney injury 10 (22), Others 10 (22) |
| Jhanji - 2010[17] - SV subgroup | Major abdominal | 45 | 5 (11) | 14 (11-26) | 26 (58) | Cardiovascular 3 (7), Infectious 24 (53), Acute kidney injury 13 (29), Others 14 (31) |
| Jhanji - 2010[17] - SV + Dopexamine subgroup | Major abdominal | 45 | 4 (9) | 16 (11-28) | 31 (69) | Cardiovascular 3 (7), Infectious 28 (62), Acute kidney injury 4 (9), Others 12 (27) |
| Suker - 2019[12] | Major abdominal | 19 | ND | ND | 6 (32) | Intra-abdominal abscess 3 (16), Pancreatic fistula 1 (5), Bile leakage 1 (5), Liver necrosis 1 (5) |

| | | | | | | |
|---|---|---|---|---|---|---|
| Dekker - 2019[7] | Cardiac Surgery | 17 | ND | 6 (5-8) | ND | ND |
| Arnold - 2012[11] | Cardiac Surgery | 20 | None | ND | None | ND |

ND, no data; CVP, central venous pressure; SV, stroke volume

**Supplementary Table 10. Risk of bias in the outcome assessment**

| Study | Method of assessment of microcirculation | Comments |
|---|---|---|
| De Baker - 2009[7]<br>Özarslan - 2012[12]<br>O'Neil - 2018[10] | Sublingual<br>Orthogonal polarization spectral imaging | Studied with a first-generation hand-held vital microscope. An externally filtered light source illuminates the organ surface with linearly polarized light, and the reflected light is blocked by an orthogonally polarized analyzer. Although these first-generation devices pioneered the initial studies, they are no longer commercially available. |

| | | |
|---|---|---|
| Jhanji - 2009[8]<br>Jhanji - 2010[17]<br>Arnold - 2012[11]<br>Bansch - 2014[13]<br>Koning - 2014[18]<br>Koning - 2016[9]<br>Dekker - 2019[7]<br>Mohamed - 2019[9]<br>Astapenko - 2019[10] | Sublingual<br>Sidestream dark-field imaging | Studied with a second-generation hand-held vital microscope. Illumination is achieved by surrounding the tip of the light guide with light-emitting diodes creating dark-field illumination. |
| Suker - 2019[12]<br>Greenwood - 2021[14] | Sublingual<br>Incident dark-field imaging | Studied with a third-generation hand-held vital microscope. It employs a new hardware platform where a high-density pixel-based imaging chip and short pulsed |

| | | |
|---|---|---|
| | | illumination source under computer control synchronizes and controls illumination and image acquisition. It also incorporates a stepping motor for quantitative focusing as well as high-resolution optics. |
